# Atherosclerotic Cardiovascular Disease and Health-Related Quality of Life Among Adults in the United States: National Health Interview Survey 2013-2017

**DOI:** 10.1101/2022.08.31.22279420

**Authors:** Kobina K. Hagan, Zulqarnain Javed, Isaac Acquah, Tamer Yahya, Javier Valero-Elizondo, Adnan A. Hyder, Elias Mossiolas, Shubham Lahan, Miguel Cainzos-Achirica, Khurram Nasir

## Abstract

**Background:** A brief health-related quality of life (HRQoL) tool with construct validity for atherosclerotic cardiovascular disease (ASCVD) may facilitate integration into healthcare delivery. We examined ASCVD-related changes in the Health and Activity Limitation Index (HALex), a generic HRQoL measure comprising perceived health and activity limitation.

**Methods:** Using data of 155,130 respondents of the National Health Interview Survey 2013-2017, we evaluated HALex scores by ASCVD (angina, heart attack, and stroke). Lower HALex scores reflected worse HRQoL and a 0.03 change was regarded as the threshold for clinical significance. Multivariable two-part models were used to assess HALex changes (β, 95%CI) associated with ASCVD overall and in sex, age, and race/ethnicity groups.

**Results:** Overall, participants with ASCVD – 6.8%, representing 15.7 million adults – had lower HALex scores (0.67) than those without ASCVD (0.87). Females, age ≥ 65 years, and non-Hispanic Blacks had the lowest HALex scores. Overall, ASCVD was associated with a substantial decrement in HALex (−0.10, [−0.10, −0.09]). Interactions between ASCVD and sex, and race/ethnicity were both significant (p < 0.001). ASCVD-associated decrement in HALex was clinically greater in: females (−0.11, [−0.12, −0.10]) than in males (−0.08, [−0.09, −0.07]); and non-Hispanic Black (−0.13, [−0.15, −0.1]) than in non-Hispanic White (−0.09, [−0.10, −0.08]). Though ASCVD impact on HALex was greater in age 18-64 years (−0.09, [−0.10, −0.08]), it was not statistically different from the elderly (−0.06, [−0.07, −0.06]).

**Conclusions:** ASCVD was consistently associated with lower HRQoL, as measured by HALex, across major demographics. HALex presents a feasible HRQoL tool to implement in healthcare.

## INTRODUCTION

Atherosclerotic cardiovascular disease (ASCVD) is associated with a high burden of debilitating symptoms and emotional stress related to decompensation and recurrent events, and invasive management. With a plenitude of life-extending treatments and a shift in healthcare quality assessment to value-based care, there is a growing call for the routine use of patient-reported outcome measures (PROMs) like health-related quality of life (HRQoL) in the clinic beyond trials (1). HRQoL not only summarizes wellbeing from a patient’s perspective, but is associated with subsequent cardiovascular events, mortality, and the patterns of healthcare utilization and expenditure (2,3).

However, the implementation of HRQoL assessment in clinical workflow is limited by factors including the lengthy surveys in commonly used research tools such as the 36-item Short Form Survey (or its shortened version, Short Form 12) and other utility-based tools (4). On the other hand, the Health and Activity Limitation Index (HALex) is a generic HRQoL measure that combines two items – perceived health and physical functioning – into a global score for wellbeing (5). It is mathematically derived and is not subject to the variations other HRQoL tools with community rated utility scores have owing to differing measuring techniques and geographic variation (6–8). With a low administrative burden, HALex may be useful in the clinic for assessing the impact of chronic conditions including ASCVD.

In this cross-sectional analysis, we described the differences in HRQoL associated with the presence of ASCVD among U.S. adults using HALex. As a convenient and reliable alternative to more detailed tools, we expected ASCVD-related decrements in HALex in the total population and within sex, age, and race/ethnicity groups.

## METHODS

### Study design and sampling

This study used the 2013-2017 National Health Interview Survey (NHIS) data. The NHIS is an annual household interview survey of the United States (U.S.) civilian, non-institutionalized population sponsored by the National Center for Health Statistics and under the auspices of the Centers for Disease Control and Prevention (9). Interviews are conducted using a complex, multistage probability design to reflect changes in the distribution of the U.S. population and produce nationally representative estimates. The NHIS has four core components – Household Composition, Family, Sample Child, and the Sample Adult – which provide information on respondents’ sociodemographic characteristics, health status and activity limitations, behavior indicators, and health care access and utilization. In addition, NHIS data are supplemented with survey weights to account for selection probabilities and non-response. Since NHIS data files are publicly available and de-identified, this study was exempt from Institutional Review Board review.

We used respondents aged ≥18 years from the Family Core, Sample Adult Core, and Person files merged and pooled over five years (2013-2017). Since NHIS data files are publicly available and de-identified, this study was exempt from the purview of Houston Methodist’s Institutional Review Board.

### Study variables

#### Atherosclerotic Cardiovascular Disease

Respondents who affirmed ever receiving a clinician diagnosis of “angina pectoris”, “heart attack (or myocardial infarction)”, “coronary heart disease”, or “stroke” were classified as having ASCVD. Respondents with no information on their ASCVD status (1,052 [0.7%]) were excluded from the analysis. All other NHIS participants included in the analysis were considered to not have ASCVD.

#### The Health and Activity Limitation Index

HALex is based on two routinely assessed health indicators included in the NHIS: perceived health status and levels of physical activity limitation (5). Responses to perceived health status are subjective and include “excellent”, “very good”, “good”, “fair”, and “poor”. The assessment of physical activity limitation is objective and the responses include: 1) not limited, 2) limited in other activities, 3) limited in major activity, 4) unable to perform a major activity, 5) unable to perform instrumental activities of daily living, and 6) unable to perform activities of daily living (personal care needs). The mathematical derivation of HALex has been described extensively (10). In brief, responses to the two items are combined in a matrix of 30 health states (**Appendix**). For persons alive, HALex scores range from 0.10 to 1.00 (11).

Two outcomes were assessed – our primary outcome, mean HALex score, and a secondary binary outcome, poor HALex performance. We categorized patients with HALex scores less than 0.84 (i.e., 20^th^ percentile of the study sample distribution) as having poor HALex (12). There are no widely established cut-off values for HALex, but the threshold for a clinically relevant impact is suggested to be 0.03 (13). We interpreted our results with this threshold. In sum, data of 155,130 participants with complete information on ASCVD and HALex, and non-zero HALex scores (i.e., alive) were used in this study.

### Covariates

Other variables included in this study were age (linear) and age group (18-64 years and ≥65 years); sex (male and female); race/ethnicity (non-Hispanic White, non-Hispanic Black, Hispanic, non-Hispanic Asian, Hispanic, and Other); educational attainment (no high school diploma, high school diploma/GED equivalent, some college, and ≥ college degree); health insurance plan (uninsured, any private plan, Medicare, Medicaid, and other plans); household income; obesity (body mass index ≥30 kg/m^2^); psychological distress within 30 days before the survey; and comorbidities, which were all self-reported. Household income categories were based on the percentage of family income relative to the federal poverty limit from the U.S. Census Bureau – high income (≥ 400%), middle income (200% to < 400%), and low income (< 200%). Psychological distress was ascertained with the Kessler-6 Distress Scale score (14). For comorbidities, respondents were asked if they had ever received a clinician diagnosis of arthritis, cancer, chronic obstructive pulmonary disease (COPD), diabetes mellitus, hypertension, or failing/weak kidneys.

### Statistical analysis

All statistical analyses incorporated the complex survey design and weighting for selection probabilities and non-response. Variance estimation for the entire pooled cohort was obtained from the Integrated Public Use Microdata Series (https://www.nhis.ipums.org) (15). We assessed the statistical significance of our estimates with a two-tailed alpha significance level of 5%. We used Stata version 16 software (StataCorp, College Station, Texas) for all analyses.

We summarized the distribution of individual characteristics – mean (SD) for age; median (IQR) for Kessler 6 score; sample frequency with weighted proportion for discrete variables) in the total population and by ASCVD status. Chi-squared, t-test, and Mann-Whitney u test statistics were used to compare the descriptive statistics between the ASCVD groups for discrete variables, linear age, and Kessler 6 score, respectively. For HALex, we provided age-and-sex adjusted least-square mean scores (with SE) for the total population and by ASCVD status.

For our primary multivariable analysis, we evaluated the associations between ASCVD and HALex scores using two-part models (16). All models used accounted for linear age, sex, race/ethnicity, insurance status, household income level, educational attainment, obesity, Kessler 6 score, and the individual comorbidities. For the two-part modeling of HALex scores, we first performed a simple negative linear transformation (X = 1 – U, where U = HALex) to move the utility index to a “health-utility decrement” scale with right skewness (16). Next, we fitted a first-part logit model for the likelihood of positive (vs. zero) scores, and an ordinary least squares regression in the second part for the predicted score conditioned on having a positive score. The same set of covariates were used in both models. Finally, a simple reconversion of the estimates and 95% confidence limits (U = 1 – X) was performed to obtain results on the original HALex scale (16). About 86% of participants had complete data on all variables of interest and the highest missingness was 7.5% for household income levels with missingness for other variables ranging from 0.8 to 3.3%. Consequently, we did not impute values for any missing data.

In subgroup analyses by age group (18-64 years and ≥ 65 years), sex, and race/ethnicity, we repeated the fully adjusted analysis of HALex scores and tested for potential interactions between these characteristics and ASCVD status separately.

In a secondary analysis of poor HALex, we used odds ratios (OR, 95% CI) to assess the association between ASCVD and poor HALex in the overall population and in subgroups defined by age, sex, and race/ethnicity.

## RESULTS

In our sample of 155,130 NHIS participants with complete HALex data, the estimated prevalence of ASCVD was 6.8%, representing about 15.7 million adults annually (**Table 1**). Individuals with ASCVD were more likely to be men, older, have Medicare insurance plan, and reside in low-income households. They also reported a greater burden of comorbidities.

**Table 1.**
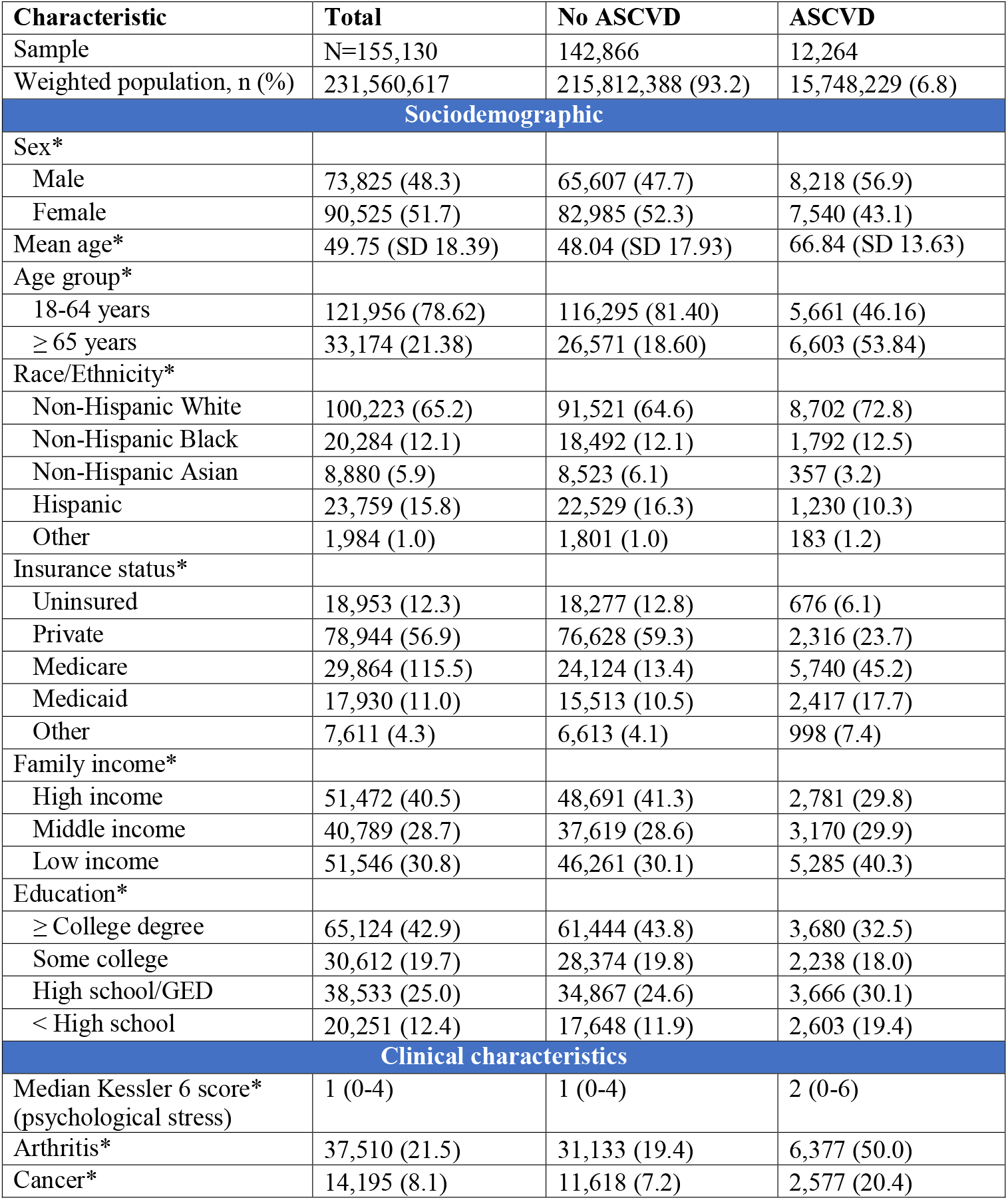
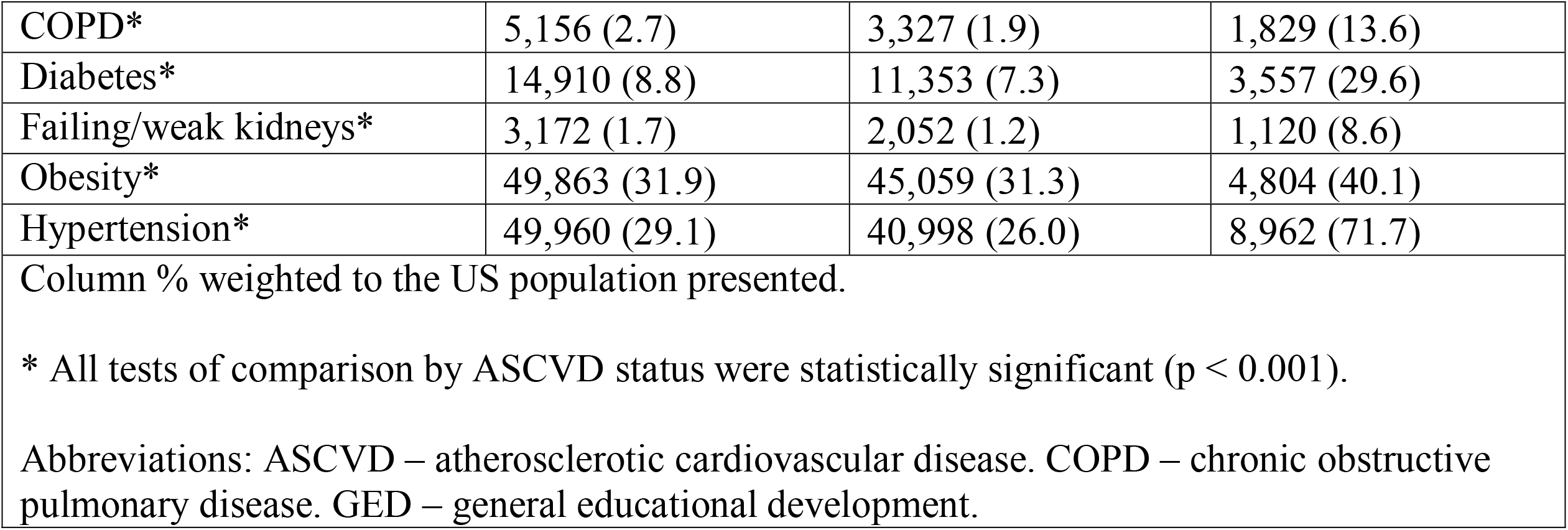
Characteristics of adults ≥ 18 years from the National Health Interview Survey (2013-2017).

### Summary of HALex scores

Overall, adults with ASCVD had lower mean HALex scores (0.67 [SE 0.01]) than those without ASCVD (0.87 [SE 0.00]). This trend was observed across all study characteristics (**Table 2)**. With ASCVD, females averaged a significantly lower HALex score than males (0.62 vs. 0.69), a contrast to the similar scores observed in the absence of ASCVD. With ASCVD, persons aged 18-64 years averaged a lower HALex score than those aged ≥ 65 years. The reverse – higher HALex scores among persons 18-64 years of age – was observed in the absence of ASCVD. Non-Hispanic Blacks averaged the lowest HALex scores, irrespective of ASCVD status. Lower educational status, lower household income and higher psychological distress were all associated with significantly lower HALex scores than the respective counterparts in both ASCVD groups. Of the comorbidities, COPD and kidney failure were associated with the lowest HALex scores reflecting their debilitating nature.

**Table 2.**
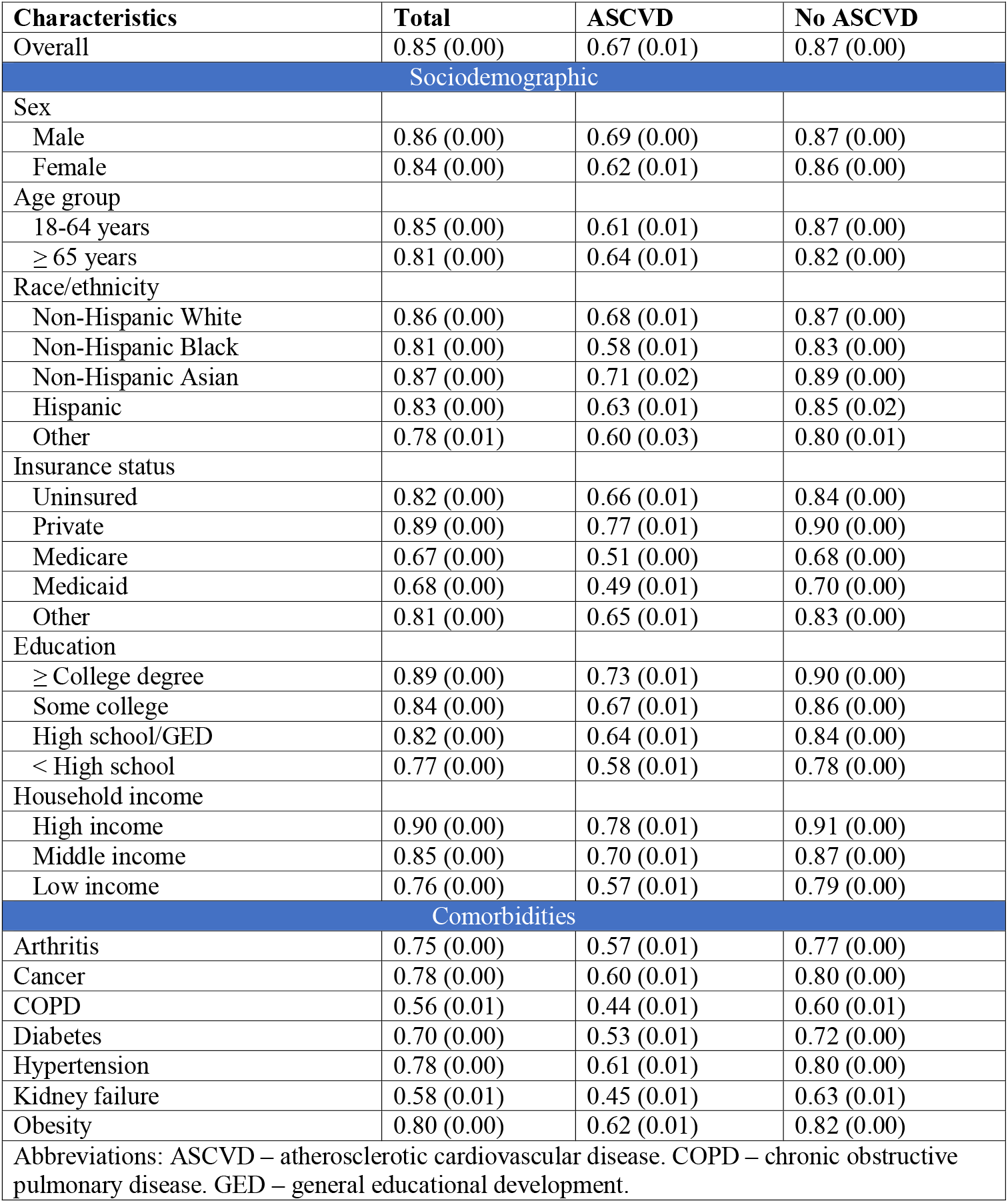
Age- and sex-adjusted mean HALex scores of adults ≥ 18 years from the National Health Interview Survey (2013-2017).

| **Characteristics** | **Total** | **ASCVD** | **No ASCVD** |
| --- | --- | --- | --- |
| Overall | 0.85 (0.00) | 0.67 (0.01) | 0.87 (0.00) |
| **Sociodemographic** |  |  |  |
| Sex |  |  |  |
| Male | 0.86 (0.00) | 0.69 (0.00) | 0.87 (0.00) |
| Female | 0.84 (0.00) | 0.62 (0.01) | 0.86 (0.00) |
| Age group |  |  |  |
| 18-64 years | 0.85 (0.00) | 0.61 (0.01) | 0.87 (0.00) |
| $\geq 65$ years | 0.81 (0.00) | 0.64 (0.01) | 0.82 (0.00) |
| Race/ethnicity |  |  |  |
| Non-Hispanic White | 0.86 (0.00) | 0.68 (0.01) | 0.87 (0.00) |
| Non-Hispanic Black | 0.81 (0.00) | 0.58 (0.01) | 0.83 (0.00) |
| Non-Hispanic Asian | 0.87 (0.00) | 0.71 (0.02) | 0.89 (0.00) |
| Hispanic | 0.83 (0.00) | 0.63 (0.01) | 0.85 (0.02) |
| Other | 0.78 (0.01) | 0.60 (0.03) | 0.80 (0.01) |
| Insurance status |  |  |  |
| Uninsured | 0.82 (0.00) | 0.66 (0.01) | 0.84 (0.00) |
| Private | 0.89 (0.00) | 0.77 (0.01) | 0.90 (0.00) |
| Medicare | 0.67 (0.00) | 0.51 (0.00) | 0.68 (0.00) |
| Medicaid | 0.68 (0.00) | 0.49 (0.01) | 0.70 (0.00) |
| Other | 0.81 (0.00) | 0.65 (0.01) | 0.83 (0.00) |
| Education |  |  |  |
| ≥ College degree | 0.89 (0.00) | 0.73 (0.01) | 0.90 (0.00) |
| Some college | 0.84 (0.00) | 0.67 (0.01) | 0.86 (0.00) |
| High school/GED | 0.82 (0.00) | 0.64 (0.01) | 0.84 (0.00) |
| $<$ High school | 0.77 (0.00) | 0.58 (0.01) | 0.78 (0.00) |
| Household income |  |  |  |
| High income | 0.90 (0.00) | 0.78 (0.01) | 0.91 (0.00) |
| Middle income | 0.85 (0.00) | 0.70 (0.01) | 0.87 (0.00) |
| Low income | 0.76 (0.00) | 0.57 (0.01) | 0.79 (0.00) |
| **Comorbidities** |  |  |  |
| Arthritis | 0.75 (0.00) | 0.57 (0.01) | 0.77 (0.00) |
| Cancer | 0.78 (0.00) | 0.60 (0.01) | 0.80 (0.00) |
| COPD | 0.56 (0.01) | 0.44 (0.01) | 0.60 (0.01) |
| Diabetes | 0.70 (0.00) | 0.53 (0.01) | 0.72 (0.00) |
| Hypertension | 0.78 (0.00) | 0.61 (0.01) | 0.80 (0.00) |
| Kidney failure | 0.58 (0.01) | 0.45 (0.01) | 0.63 (0.01) |
| Obesity | 0.80 (0.00) | 0.62 (0.01) | 0.82 (0.00) |
| Abbreviations: ASCVD – atherosclerotic cardiovascular disease. COPD – chronic obstructive pulmonary disease. GED – general educational development. |  |  |  |

### Multivariable ASCVD-HALex score analysis

We present our multivariable analysis of HALex scores. Overall, ASCVD was associated with a clinically relevant decrease in HALex performance (β = −0.10; 95% CI [−0.10, −0.09]) (**Figure 1**). We observed, in separate models, significant interactions between ASCVD and sex (p < 0.001), and race/ethnicity (p < 0.001). The decrement in HALex scores associated with ASCVD was greater in females (β = −0.11, 95% CI [−0.12, −0.10]) than in males (β = −0.08, 95% CI [−0.09, - 0.07]). For age groups, the negative impact of ASCVD on HALex scores was observed to be greater in the younger age group (β = −0.09, 95% CI [−0.10, −0.08]) than those aged ≥ 65 years (β = −0.06, 95% CI [−0.07, −0.06]). For racial/ethnic subgroups, the greatest ASCVD impact was observed with non-Hispanic Blacks (β = −0.13, 95% CI [−0.15, −0.11]). The decrement in HALex associated with ASCVD was not clinically different between non-Hispanic White (β = −0.09, 95% CI [−0.10, −0.08]) and Hispanics (β = −0.10, 95% CI [−0.12, −0.08]) groups.

**Figure 1.**
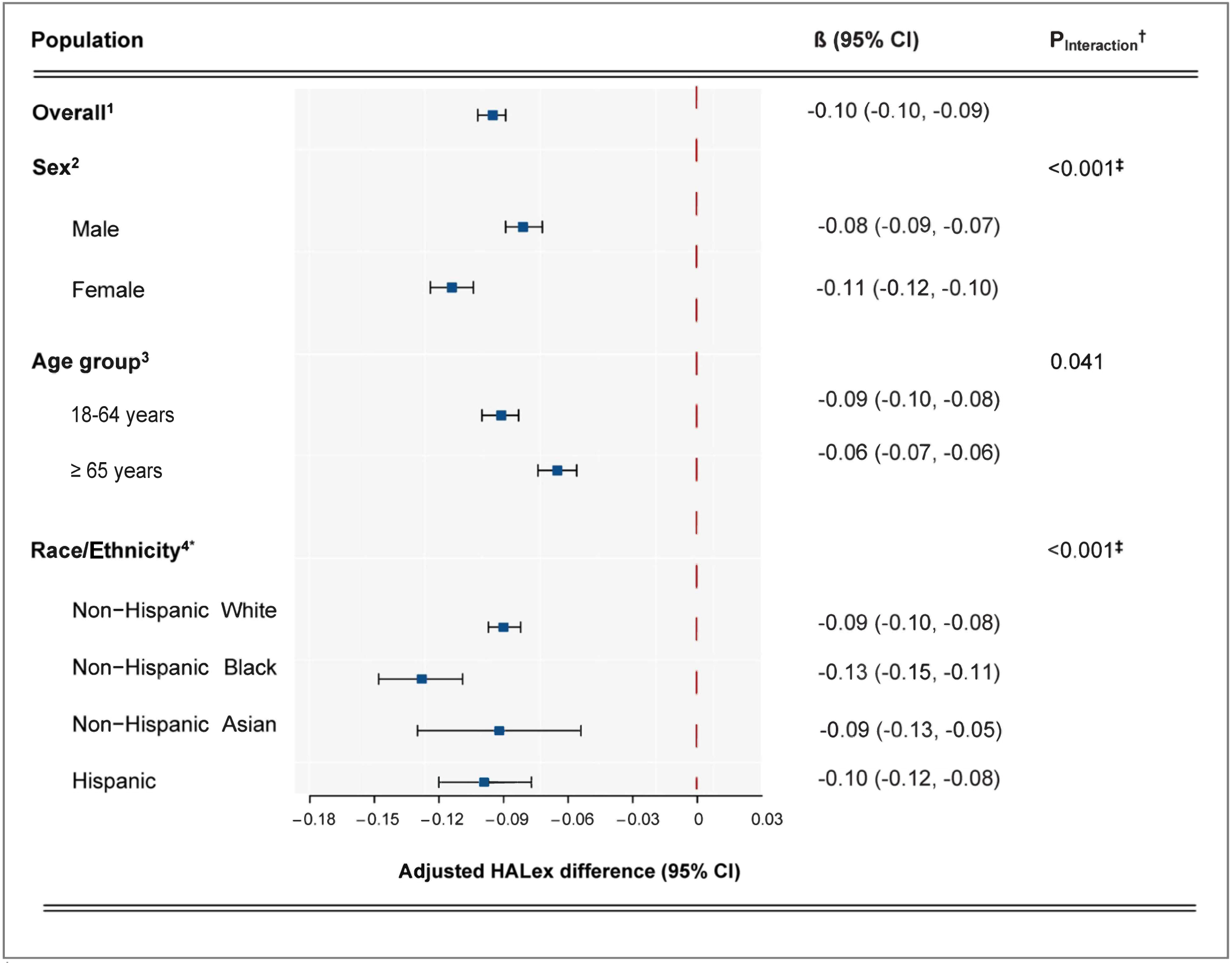
Association between HALex and ASCVD from the National Health Interview Survey 2013-2017. [Caption] ^1^ Adjusted for sex, age (linear), race/ethnicity, insurance status, household income level, educational attainment, obesity, Kessler-6 score, and comorbidities. ^2^ Adjusted for age (linear), race/ethnicity, insurance status, household income level, educational attainment, obesity, Kessler-6 score, and comorbidities. ^3^ Adjusted for sex, race/ethnicity, insurance status, household income level, educational attainment, obesity, Kessler-6 score, and comorbidities. ^4^ Adjusted for sex, age (linear), insurance status, household income level, educational attainment, obesity, psychological distress, and comorbidities. * *Other* race/ethnicity group not shown due to limited sample size. ^†^ p-value for the test of interaction with Bonferroni correction. ^‡^ Statistically significant.

### Secondary analysis of poor HALex

Overall, 17.7% (95% CI [17.4, 18.1]) of the study population performed poorly on HALex. A significantly greater proportion in the ASCVD group had poor HALex than those without ASCVD (40.3% v. 15.3%; p < 0.001; **Appendix**). The distribution of poor HALex performance across study variables for the ASCVD groups are presented as well in the Appendix.

From the multivariable analysis, ASCVD was associated with increased odds of poor HALex 2.5 times that observed with the group without ASCVD (OR = 2.46, 95% CI [2.28, 2.66]). Interactions between ASCVD and age group and ASCVD and race/ethnicity were statistically significant (p < 0.001), but there was no significant interaction observed between ASCVD and sex (p = 0.05). The increment in odds of poor HALex associated with ASCVD was higher in age 18-64 years (OR = 2.84; 95% CI [2.52, 3.20]) than in age ≥65 years. Similarly, the increment in odds of poor HALex associated with ASCVD was highest in the non-Hispanic Black group (OR = 2.33; 95% CI [2.74, 4.04]). Hispanic and non-Hispanic White groups had similar increments in odds of poor HALex associated with ASCVD presence.

## DISCUSSION

In a large survey of adults in the U.S., reporting any ASCVD condition was associated with a significantly lower HRQoL (as measured with HALex). This clinically significant difference was independent of demography, education, insurance, household income, and comorbidity burden. Further, the impact of ASCVD on lower HALex scores was significantly greater in females, adults 18-64 years of age, and non-Hispanic Black adults. The impacts of ASCVD on HALex scores and poor HALex performance were similar in the Hispanic and non-Hispanic White groups. We did not intend to establish clinical values of HALex for persons with ASCVD. Rather, we set out to corroborate the construct validity of a HRQoL tool that could be implemented in the clinical workflow without much challenge.

The sex differences in the impact of ASCVD on HALex is congruent with the established trend that women have poorer health outcomes in conditions like coronary heart disease (17). Women may present as a diagnostic dilemma in the clinic due to confounding atypical presentations or less common pathophysiological processes. Consequently, they are less likely to receive prompt and/or appropriate management to realize optimal outcomes (18,19).

We also observed worse HRQoL and the largest ASCVD decrement in non-Hispanic Black compared to the other groups. A similar trend was previously observed in a national survey of adults 35-89 years of age which assessed HRQoL with various indices (20). Interestingly, the impact of ASCVD on HRQoL in Hispanic was comparable to non-Hispanic White, despite the former averaging lower HALex in the presence of ASCVD. This paradox, commonly observed in population studies on other health outcomes (21), could be related to a more resilient subjective construct of health in the face of health challenges owing to cultural values and supportive social relationships among Hispanics (22).

On average, we observed lower HALex scores in those 18-64 years of age with ASCVD than their older counterparts. Though not statistically significant, we also observed a greater decrement in HALex with ASCVD in the group 18-64 years of age (−0.09 vs −0.06). While younger age is associated with better health and a greater chance of recovering from ASCVD and its management, younger individuals are more likely to have maladaptive and adverse psychological experiences after their diagnosis (23). Another potential explanation to this observation is the phenomenon that aging populations may report better subjective health due to lowered expectations rather than ‘actual’ better health (24).

### Implications

The adoption into clinical workflow of a HRQoL tool like HALex – which is easy to administer and has construct validity – would be in line with the call to routinely assess PROMs in the clinic. HALex is impacted by the burden of chronic disease, more so than other preference-based measures (25,26). This is in part due to the ability of the constituent perceived-health-status item to discriminate well among levels of functioning (27). As physical functioning is strongly associated with cardiovascular disease prognosis, HALex assessment could be relevant in patient management, especially in the holistic evaluation of rehabilitation (28).

Additionally, HALex may have utility in longitudinally assessing the impact of treatment on HRQoL (29). Under a chronic care model with self-management support strategy, patients are reinforced to assume an active and shared responsibility of their health with providers. They are educated to better recognize their health issues, acknowledge the need for health behavior modifications, and initiate and maintain such modifications. In utilizing this context of care, perceived health status (and HALex) would be based on a more comprehensive conceptualization of health standards set and evaluated by patients themselves. Consequently, for patients who may not have a change in physical functioning, a change in their valuation of their own health could signal deficiencies in care standards.

### Limitations

HALex as a HRQoL measure is not without limitations. Firstly, domains of health such as emotional, mental, and social functioning, which are relevant in HRQoL (30), are omitted in HALex derivation. This may its discrimination ability, especially for populations with clustering at the highest level of health (5). Nevertheless, we accounted for psychological distress using a valid and reliable tool in the Kessler 6 distress scale, which minimizes the bias from this health domain on our estimates.

Secondly, the reliance of HALex on a subjectively perceived health status raises the question of how much of the difference in HALex scores may be related to differential perception. Although perceived health status tends to be congruous with objective measures of health, differences exist between the two health assessments owing to varying health expectations (or preoccupations), and the relevance of physical function to major lifetime occupation (31). Nevertheless, it is well known that the manner in which people account for the many dimensions of health when rating their overall health is relatively stable across disease populations.

Finally, while the NHIS is designed to be representative of the US population, the reliance on self-reports for health information including medical conditions and anthropometric measurements (i.e., weight and height) implies some potential for misclassification of study variables. However, previous studies have found a high correlation between the self-reported information in NHIS and the verified information found in other national datasets (32).

## CONCLUSION

In conclusion, ASCVD is associated with lower HALex and this association was observed to be greater in female adults, younger age group, and non-Hispanic Black persons. HALex retains a construct validity and has a low administration burden, making it a potentially useful HRQoL tool to implement in healthcare delivery.

## Data Availability

All data produced in the present study are available upon reasonable request to the authors

https://www.cdc.gov/nchs/nhis/1997-2018.htm

## Non-standard Abbreviations and Acronyms

ASCVD: atherosclerotic cardiovascular disease
COPD: chronic obstructive pulmonary disease
GED: General Educational Development
HALex: Health and Activity Limitation Index
HRQoL: health-related quality of life
NHIS: National Health Interview Survey
OR: odds ratio
PROM: patient-reported outcome measures
SE: standard error

## DECLARATIONS

### Ethics approval and consent to participate

Not applicable.

### Consent for publication

Not applicable.

### Availability of data and materials

The datasets analyzed in the current study are available from the corresponding author on reasonable request.

### Authors’ contributions

K.K.H., K.N., and Z.J. contributed to the study’s conception and design. Material preparation and data analysis were performed by K.K.H. and J.VE. The first draft of the manuscript was prepared by K.K.H. and Z.J. S.L. prepared all figures. All authors reviewed the manuscript. All authors read and approved the final manuscript.

## Acknowledgements

Not applicable.

