## Appendix for "Atherosclerotic Cardiovascular Disease and Health-Related Quality of Life Among Adults in the United States: National Health Interview Survey 2013-2017"

**SUPPLEMENTAL MATERIAL**

**SUPPLEMENTAL APPENDIX**

1. **Supplemental Table I**. Values assigned to health states defined by perceived health and activity limitation (HALex scores)

2. **Supplemental Table II.** Sex differences in HALex values by ASCVD status across sociodemographic categories

3. **Supplemental Table III.** Beta-coefficients of covariates of interest for predicted HALex scores

4. **Supplemental Figure I.** Distribution of Health and Activity Limitation Index

**Supplemental Table I.** Values assigned to health states defined by perceived health and activity limitation (HALex scores)

|  |  | Perceived health status | | | | | |
| --- | --- | --- | --- | --- | --- | --- | --- |
|  |  | Excellent | Very good | Good | Fair | Poor | Dead |
| Activity limitation | Single attributable score | 1.00 | 0.85 | 0.70 | 0.30 | 0.00 |  |
| Not limited | 1.00 | 1.00 | 0.92 | 0.84 | 0.63 | 0.47 |  |
| Limited in performing other activities | 0.75 | 0.87 | 0.79 | 0.72 | 0.52 | 0.38 |  |
| Limited in performing major activities | 0.65 | 0.81 | 0.74 | 0.67 | 0.48 | 0.34 |  |
| Unable to perform major activity | 0.40 | 0.68 | 0.62 | 0.55 | 0.38 | 0.25 |  |
| Limited in instrumental activities of daily living (IADL) | 0.20 | 0.57 | 0.51 | 0.45 | 0.29 | 0.17 |  |
| Limited in activities of daily living (ADL) | 0.00 | 0.47 | 0.41 | 0.36 | 0.21 | 0.10 |  |
| Dead |  |  |  |  |  |  | 0.00 |

**Supplemental Table II.** Distribution of HALex scores by ASCVD status

| ASCVD | | | | | | |
| --- | --- | --- | --- | --- | --- | --- |
| Activity limitation | Perceived health status | | | | |  |
|  | Excellent | Very good | Good | Fair | Poor | Total |
| Not limited | 5.95% | 16.9% | 22.5% | 9.8% | 1.6% | 56.8% |
| Limited other | 0.1% | 0.6% | 1.8% | 1.5% | 0.2% | 4.2% |
| Limited major | 0.0% | 0.3% | 1.6% | 1.5% | 0.6% | 4.1% |
| Unable major | 0.0% | 0.5% | 2.2% | 5.3% | 3.2% | 11.3% |
| Limited in IADL | 0.1% | 0.7% | 2.6% | 4.4% | 3.0% | 10.9% |
| Limited in ADL | 0.0% | 0.4% | 2.1% | 4.6% | 5.5% | 12.7% |
| Total | 6.5% | 19.3% | 32.9% | 27.1% | 14.2% | 100% |
| No ASCVD | | | | | | |
| Activity limitation | Perceived health status | | | | |  |
|  | Excellent | Very good | Good | Fair | Poor | Total |
| Not limited | 30.4% | 32.5% | 22.1% | 4.3% | 0.3% | 89.6% |
| Limited other | 0.1% | 0.4% | 0.5% | 0.3% | 0.0% | 1.4% |
| Limited major | 0.2% | 0.5% | 0.9% | 0.5% | 0.1% | 2.1% |
| Unable major | 0.1% | 0.3% | 1.1% | 1.3% | 0.6% | 3.4% |
| Limited in IADL | 0.0% | 0.2% | 0.6% | 0.7% | 0.4% | 2.0% |
| Limited in ADL | 0.0% | 0.1% | 0.3% | 0.6% | 0.5% | 1.6% |
| Total | 31.0% | 34.0% | 25.5% | 7.7% | 1.9% | 100% |

**Supplemental Table III.** Gender differences in HALex values by ASCVD status across sociodemographic categories

| Characteristics | ASCVD | | | No ASCVD | | |
| --- | --- | --- | --- | --- | --- | --- |
|  | Men | Women | Difference | Men | Women | Difference |
| Age strata |  |  |  |  |  |  |
| 18-39 | 0.71 (0.01) | 0.72 (0.01) | -0.00 (0.03) | 0.91 (0.00) | 0.90 (0.00) | 0.01 (0.00)* |
| 40-54 | 0.62 (0.01) | 0.59 (0.01) | 0.03 (0.02)* | 0.86 (0.00) | 0.85 (0.00) | 0.01 (0.00)* |
| 55-64 | 0.61 (0.01) | 0.55 (0.01) | 0.05 (0.01)* | 0.82 (0.00) | 0.82 (0.00) | 0.01 (0.00) |
| 65+ | 0.71 (0.01) | 0.58 (0.01) | 0.13 (0.01)* | 0.84 (0.00) | 0.81 (0.00) | 0.03 (0.00)* |
| Race/ethnicity |  |  |  |  |  |  |
| NH White | 0.68 (0.01) | 0.61 (0.01) | 0.07 (0.01)* | 0.88 (0.00) | 0.86 (0.00) | 0.01 (0.00)* |
| NH Black | 0.57 (0.01) | 0.51 (0.01) | 0.06 (0.02)* | 0.85 (0.00) | 0.83 (0.00) | 0.02 (0.00)* |
| NH Asian | 0.72 (0.01) | 0.59 (0.01) | 0.13 (0.04)* | 0.90 (0.00) | 0.89 (0.00) | 0.01 (0.00)* |
| Hispanic | 0.62 (0.01) | 0.57 (0.01) | 0.04 (0.02) | 0.88 (0.00) | 0.86 (0.00) | 0.02 (0.00)* |
| Insurance status |  |  |  |  |  |  |
| Uninsured | 0.69 (0.01) | 0.62 (0.01) | 0.08 (0.03)* | 0.87 (0.00) | 0.86 (0.00) | 0.02 (0.00)* |
| Private | 0.76 (0.01) | 0.76 (0.01) | 0.00 (0.01) | 0.91 (0.00) | 0.90 (0.00) | 0.01 (0.00)* |
| Medicare | 0.68 (0.01) | 0.58 (0.01) | 0.10 (0.01)* | 0.80 (0.00) | 0.79 (0.00) | 0.01 (0.00) |
| Medicaid | 0.45 (0.01) | 0.41 (0.01) | 0.04 (0.01)* | 0.73 (0.00) | 0.75 (0.00) | -0.02 (0.01)* |
| Other | 0.62 (0.01) | 0.58 (0.02) | 0.04 (0.03) | 0.81 (0.00) | 0.83 (0.01) | -0.02 (0.01)* |
| Education |  |  |  |  |  |  |
| College+ | 0.73 (0.01) | 0.66 (0.01) | 0.07 (0.01)* | 0.91 (0.00) | 0.90 (0.00) | 0.01 (0.00)* |
| Some college | 0.66 (0.01) | 0.61 (0.01) | 0.05 (0.02)* | 0.88 (0.00) | 0.86 (0.00) | 0.02 (0.00)* |
| High school/GED | 0.64 (0.01) | 0.58 (0.01) | 0.06 (0.01)* | 0.85 (0.00) | 0.83 (0.00) | 0.02 (0.00)* |
| < High school | 0.55 (0.01) | 0.47 (0.01) | 0.08 (0.01)* | 0.81 (0.00) | 0.77 (0.00) | 0.04 (0.00)* |
| Family income |  |  |  |  |  |  |
| High-income | 0.78 (0.01) | 0.71 (0.01) | 0.07 (0.01)* | 0.91 (0.00) | 0.91 (0.00) | 0.01 (0.00)* |
| Middle-income | 0.67 (0.01) | 0.64 (0.01) | 0.04 (0.01)* | 0.88 (0.00) | 0.86 (0.00) | 0.01 (0.00)* |
| Low-income | 0.58 (0.01) | 0.55 (0.01) | 0.04 (0.02)* | 0.83 (0.00) | 0.82 (0.00) | 0.01 (0.00)* |
| Lowest-income | 0.48 (0.01) | 0.45 (0.01) | 0.03 (0.01)* | 0.80 (0.00) | 0.78 (0.00) | 0.02 (0.00)* |

3. **Supplemental Table IV.** Beta-coefficients of covariates of interest for predicted HALex scores

| Covariate | Unadjusted β-coefficient | Adjusted β-coefficient^*^ | |
| --- | --- | --- | --- |
|  |  | ASCVD | No ASCVD |
| Gender |  |  |  |
| Men | Reference | Reference | Reference |
| Women | -0.01 (-0.02, -0.01)^†^ | -0.03 (-0.04, -0.01)^†^ | 0.00 (-0.00, 0.00) |
| Age strata, years |  |  |  |
| 18-39 | Reference | Reference | Reference |
| 40-54 | -0.05 (-0.05, -0.04)^†^ | -0.06 (-0.09, -0.03)^†^ | -0.03 (-0.04, -0.03) |
| 55-64 | -0.08 (-0.09, -0.08)^†^ | -0.07 (-0.10, -0.05)^†^ | -0.05 (-0.05, -0.04) |
| 65+ | -0.08 (-0.09, -0.08)^†^ | 0.05 (0.02, 0.08)^†^ | 0.05 (0.05, 0.06) |
| Race/ethnicity |  |  |  |
| Non-Hispanic White | Reference | Reference | Reference |
| Non-Hispanic Black | -0.03 (-0.04, -0.03)^†^ | -0.03 (-0.05, -0.01)^†^ | -0.00 (-0.01, 0.00) |
| Non-Hispanic Asian | 0.03 (0.03, 0.04)^†^ | 0.00 (-0.03, 0.03) | -0.00 (-0.01, 0.00) |
| Hispanic | -0.00 (-0.00, 0.01)^†^ | 0.01 (-0.02, 0.03) | 0.01 (0.01, 0.01)^†^ |
| Insurance |  |  |  |
| Private | Reference | Reference | Reference |
| Uninsured | -0.04 (-0.05, -0.04)^†^ | -0.03 (-0.05, -0.00)^†^ | -0.01 (-0.02, -0.01)^†^ |
| Medicare | -0.14 (-0.15, -0.14)^†^ | -0.15 (-0.17, -0.13)^†^ | -0.11 (-0.12, -0.11)^†^ |
| Medicaid | -0.20 (-0.20, -0.19)^†^ | -0.21 (-0.24, -0.18)^†^ | -0.10 (-0.10, -0.09)^†^ |
| Household income |  |  |  |
| High income | Reference | Reference | Reference |
| Middle income | -0.05 (-0.05, -0.04)^†^ | -0.04 (-0.06, -0.02)^†^ | -0.02 (-0.03, -0.02)^†^ |
| Low income | -0.10 (-0.10, -0.09)^†^ | -0.08 (-0.10, -0.06)^†^ | -0.04 (-0.05, -0.04)^†^ |
| Lowest income | -0.14 (-0.15, -0.14)^†^ | -0.12 (-0.15, -0.10)^†^ | -0.06 (-0.07, -0.06)^†^ |
| Education |  |  |  |
| College+ | Reference | Reference | Reference |
| Some college | -0.04 (-0.04, -0.03)^†^ | -0.02 (-0.04, -0.00)^†^ | -0.01 (-0.01, -0.01)^†^ |
| High school/GED | -0.07 (-0.07, -0.07)^†^ | -0.03 (-0.05, -0.02)^†^ | -0.02 (-0.02, -0.01)^†^ |
| < High school | -0.13 (-0.14, -0.13)^†^ | -0.08 (-0.10, -0.06)^†^ | -0.05 (-0.05, -0.04)^†^ |
| Smoking history |  |  |  |
| Never | Reference | Reference | Reference |
| Former | -0.05 (-0.05, -0.05)^†^ | -0.01 (-0.00, -0.02)^†^ | -0.01 (-0.01, -0.01)^†^ |
| Current | -0.08 (-0.09, -0.08)^†^ | -0.02 (-0.04, -0.00)^†^ | -0.03 (-0.04, -0.03) ^†^ |
| Arthritis | -0.14 (-0.15, -0.14)^†^ | -0.07 (-0.08, -0.05)^†^ | -0.07 (-0.07, -0.07)^†^ |
| Cancer | -0.09 (-0.10, -0.09)^†^ | -0.03 (-0.04, -0.01)^†^ | -0.04 (-0.04, -0.03)^†^ |
| COPD | -0.29 (-0.30, -0.28)^†^ | -0.12 (-0.14, -0.10)^†^ | -0.15 (-0.16, -0.14)^†^ |
| Diabetes or prediabetes | -0.19 (-0.19, -0.18)^†^ | -0.08 (-0.10, -0.06)^†^ | -0.09 (-0.09, -0.08)^†^ |
| Obesity | -0.07 (-0.07, -0.07)^†^ | -0.02 (-0.04, -0.01)^†^ | -0.03 (-0.04, -0.03)^†^ |
| Kidney failure | -0.29 (-0.31, -0.28)^†^ | -0.14 (-0.17, -0.11)^†^ | -0.13 (-0.15, -0.12)^†^ |
| ^*^ Model adjusted for age, sex, race/ethnicity, insurance, household income, educational attainment, smoking history, arthritis, cancer, COPD, diabetes/prediabetes, kidney failure, and obesity.  ^†^ p-value < 0.05  Abbreviations: ASCVD – atherosclerotic cardiovascular disease. COPD – chronic obstructive pulmonary disease. GED – general educational development. | | | |

**Supplemental Figure I.** Distribution of Health and Activity Limitation Index

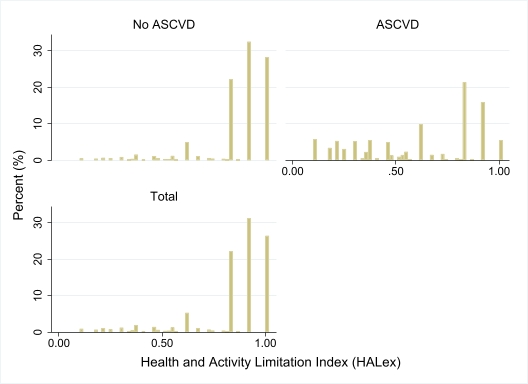
